# Quarantine, relaxation and mutation explaining the CoViD-19 epidemic in São Paulo State (Brazil)

**DOI:** 10.1101/2021.04.12.21255325

**Authors:** Hyun Mo Yang, Luis Pedro Lombardi Junior, Fabio Fernandes Morato Castro, Ariana Campos Yang

## Abstract

**Background:** The continuous SARS-CoV-2 transmission in several countries could contribute to the mutations’ appearance. The circulation of more virulent variants may increase the number of severe CoViD-19 needing hospital care and fatalities hugely.

**Methods:** The partial quarantine in São Paulo State and further relaxation associated with the mutations are explained by a mathematical model based on the CoViD-19 natural history encompassing the age-dependent fatality. The model parameters were fitted considering the observed data from São Paulo State.

**Results:** The partial quarantine was explained by the less virulent SARS-CoV-2 transmission, but the relaxation alone could not explain the epidemic observed in São Paulo State. However, more virulent variants plus the transmission among isolated individuals explained the increased CoViD-19 fatalities.

**Conclusions:** The model described the CoViD-19 epidemic in São Paulo State by considering the partial quarantine, relaxation and mutations. The model provided a potential epidemiological scenario in the absence of mass vaccination.

## 1 Introduction

Many countries adopted partial or rigid quarantine to control the coronavirus disease 2019 (CoViD-19) pandemic caused by the severe acute respiratory syndrome coronavirus 2 (SARS-CoV-2) infection. For instance, Spain [1] and Italy [2] adopted rigid quarantine (lockdown), while Brazil (São Paulo State also) [3] adopted the partial quarantine (isolation). SARS-CoV-2, an RNA virus, mutates faster, and variants of the original virus may appear. The non-pharmaceutical interventions called suppression (lockdown) and mitigation (isolation) affect the SARS-CoV-2 transmission differently and, consequently, may influence mutant variants’ appearance. Especially the variants B.1.1.7 (United Kingdom), B.1.351 (South Africa), and P.1 (Brazil) challenge the epidemic’s control by vaccination [4].

In São Paulo State, the first CoViD-19 case was observed on February 26, the first death on March 16, and the partial isolation was implemented on March 24, 2020. At the end of June 2020, just after the peak of the epidemic under isolation [5], the relaxation was initiated but not fully implemented. The continuous transmission of SARS-CoV-2 may result in the appearance of more virulent variants, which explains the increased CoViD-19 fatalities compared with the epidemic with less virulent ones [6]. São Paulo State associated the variant P.1 with increased mortality in the CoViD-19 epidemic, as observed in the Amazonas State [7].

Based on a mathematical model, we evaluate the partial quarantine and further relaxation in São Paulo State. The original SARS-CoV-2 transmission fully described the isolation; however, the relaxation could not describe the CoViD-19 data collection from São Paulo State alone. We allowed the transmission of more virulent variants and the transmission among isolated individuals in the model to describe the observed CoViD-19 fatalities fully. We estimate the model parameters against the observed data from February 26, 2020, to April 5, 2021, to retrieve the CoViD-19 cases and fatalities curves. Besides fitting the epidemiological curves, we tested the model’s capability to predict the outcoming epidemic.

## 2 Material and methods

The SARS-CoV-2 transmission model encompassing young (*y*) and elder (*o*) subpopulations is described by the system of ordinary differential equations presented in Appendix A, named the SQEAPMDR model. We drop out the pulses in equation (A.2) in Appendix A and transfer them to the boundary conditions. Hence, equations for susceptible and isolated individuals are

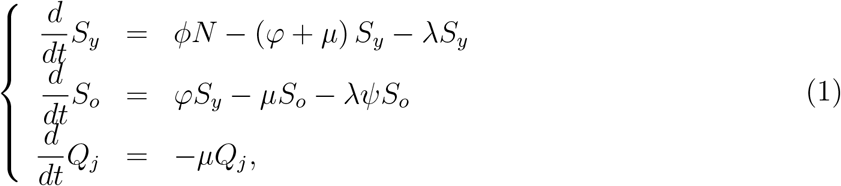

for infected individuals, with *j* = *y, o*,

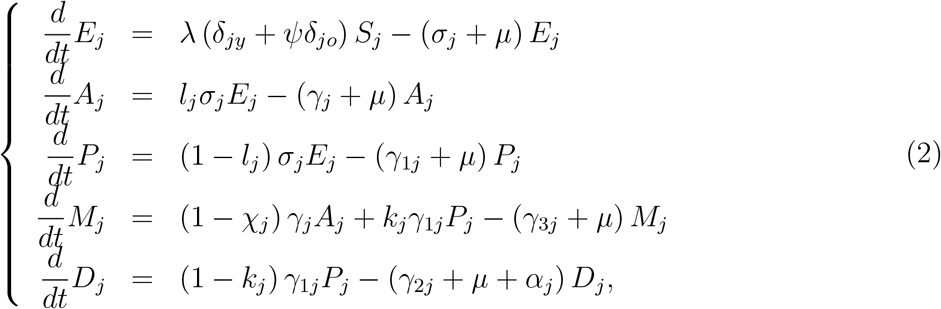

and for recovered individuals,

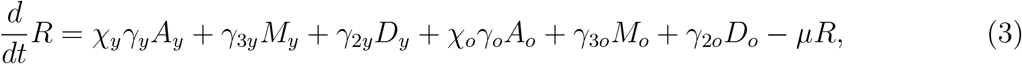

where *λ* is the force of infection given by equation (A.1), and *N*_*j*_ = *S*_*j*_ + *Q*_*j*_ + *E*_*j*_ + *A*_*j*_ + *P*_*j*_ + *M*_*j*_ + *D*_*j*_, with *N* = *N*_*y*_ + *N*_*o*_ + *R* obeying

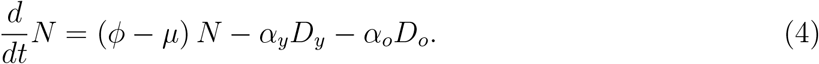

The initial number of population is *N*(0) = *N*_0_ = *N*_0*y*_ +*N*_0*o*_, where *N*_0*y*_ and *N*_0*o*_ are the size of young and elder subpopulations at *t* = 0 (calendar time February 26, 2020).

To estimate the transmission rates *β*_1*j*_, *β*_2*j*_, and *β*_3*j*_, *j* = *y, o*, the proportion in isolation *u*, the protective factor *ε*, the reduction factor *ω*, and the proportion of the *i*-th relaxation *u*_*i*_ (*x* stands for one of these parameters), we calculate

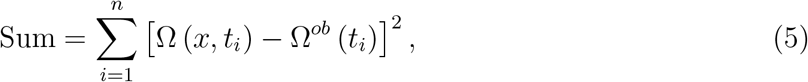

where Ω^*ob*^ (*t*_*i*_) is the accumulated severe CoViD-19 registered cases at day *t*_*i*_, and Ω (*x, t*_*i*_) is the accumulated severe CoViD-19 cases given by

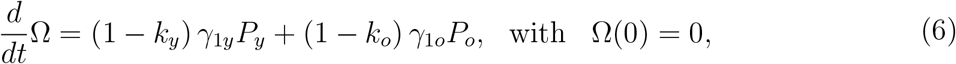

which are the exit from class *P*, and entering into class *D*. To evaluate Ω, the numerical solutions of equations (1), (2) and (3) are obtained using the initial conditions given by equation (A.3) in Appendix A. We search for the value of *x* minimizing the Sum.

To estimate the additional mortality (fatality) rates *α*_*y*_ and *α*_*o*_, we calculate

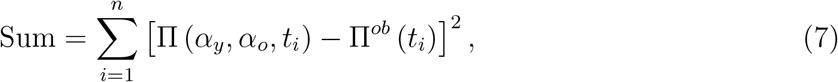

where Π^*ob*^ (*t*_*i*_) is the accumulated CoViD-19 deaths registered at day *t*_*i*_, and Π (*α*_*y*_, *α*_*o*_, *t*_*i*_) = Π_*y*_ (*α*_*y*_, *t*_*i*_) + Π_*o*_ (*α*_*o*_, *t*_*i*_) is given by

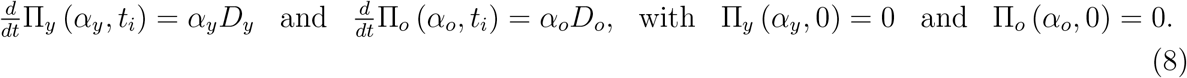

We search for the values of *α*_*y*_ and *α*_*o*_ minimizing the Sum. In the estimation of the additional mortality rates, we must bear in mind that the time at which new cases and deaths were registered does not have direct correspondence; instead, they are delayed by Δ days, that is, 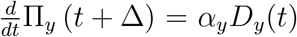 and 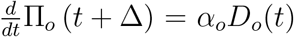. Among exits from compartment *D* (*γ*_2*j*_, *µ*, and *α*_*j*_), we are counting only the deaths caused by severe CoViD-19.

From the accumulated CoViD-19 cases and deaths estimated curves, we retrieve the daily cases and deaths. The daily CoViD-19 cases Ω_*d*_ is, considering Δ*t* = *t*_*i*_ − *t*_*i*−1_ = Δ*t* = 1 *day*,

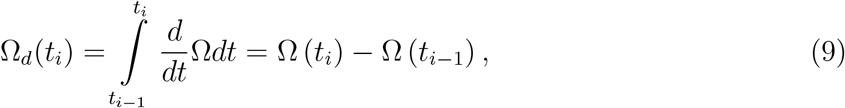

where Ω_*d*_(0) = Ω_*d*0_ is the first observed CoViD-19 case at *t* = 0, with *i* = 1, 2, …, and *t*_1_ = 1 is the next day in the calendar time, and so on. The daily CoViD-19 deaths Π_*d*_ is

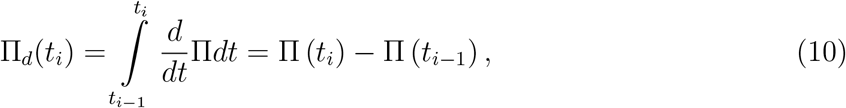

where Π_*d*_(*t*_0_) = Π_*d*0_ is the first observed CoViD-19 death at *t*_0_ (calendar time March 16, 2020). The steady-state of the system of equations in terms of fractions corresponding to equations (1), (2) and (3) can be evaluated [3]. From the stability analysis of the trivial equilibrium point, we obtain the basic reproduction number *R*_0_, which is given by equation (A.6) in Appendix A. The fractions written as 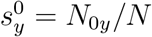 and 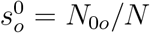 result in

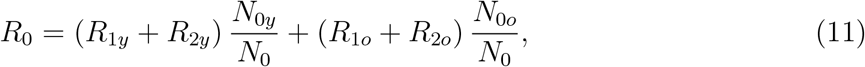

where *N*_0*y*_ and *N*_0*o*_ are the initial numbers of young and elder subpopulations with *N*_0_ = *N*_0*y*_ + *N*_0*o*_, and

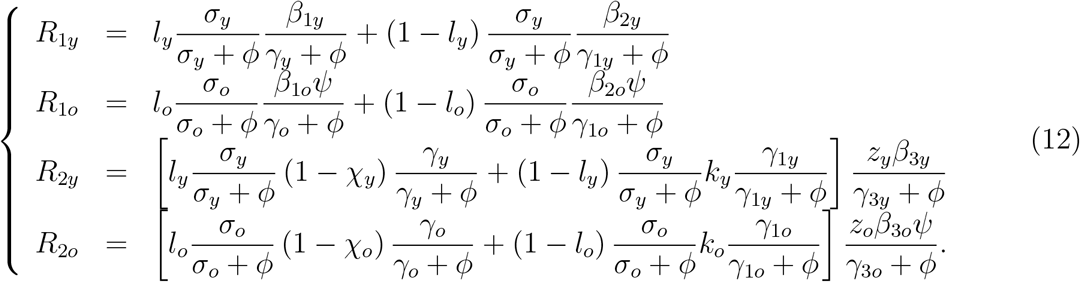

Letting *z*_*y*_ = *z*_*o*_ = 0 (*R*_2*y*_ = *R*_2*o*_ = 0), we retrieve the basic reproduction number obtained in [3].

More virulent SARS-CoV-2 enhances the virus’s capacity to infect target cells, resulting in an increased number of infected cells. Consequently, the possibility of manifesting mild or severe disease increases, increasing the symptomatic CoViD-19 cases. For this reason, we translate the virulence with the ratio between asymptomatic and symptomatic individuals (parameters *l*_*y*_ and *l*_*o*_). Another effect of higher virulence is the increased amount of virus released by infectious individuals in the environment, which must increase the transmission [8]. However, the isolation of severe and part of mild CoViD-19 cases decreases the transmission populationally [6].

## 3 Results

The CoViD-19 epidemic in São Paulo State is split into four phases, non-mutually exclusive, considering the mutation in the original SARS-CoV-2. Phase 1 is the natural epidemic, just before introducing partial quarantine; phase 2 is the epidemic under isolation; phase 3 is the epidemic under relaxation; and phase 4 is the epidemic with more virulent variant SARS-CoV-2. In the initial phases, the infections by a pool of original and less virulent variants SARS-CoV-2 prevail, and in the last phases, the more virulent variants contribute to increasing the deaths. Still, the level of transmission is assumed not to vary.

At the beginning of the epidemic, we have two data sets: Severe CoViD-19 cases (those in hospitals were tested and confirmed) and deaths. Due to the lack of mass testing (PCR and serology) when SARS-CoV-2 transmission began, the severe CoViD-19 curve *D* can be considered the epidemic curve. Hence, we estimate Ω using equation (5) and retrieve the epidemic curve *D*. From this estimated *D*, we estimate the fatality rates *α*_*y*_ and *α*_*o*_ using equation (7). However, as the epidemic evolved and more tests became available, asymptomatic and mild CoViD-19 were recorded together with the severe cases, and the recorded data can not be matched with the severe CoViD-19 epidemic curve *D* anymore. Notwithstanding, the number of fatalities is reliable data to be matched with the severe CoViD-19 cases. For this reason, we estimate the CoViD-19 deaths curve using equation (7) and indirectly retrieve the CoViD-19 epidemic curve *D*, which situates below the CoViD-19 registered data.

Phase 1 in the São Paulo State CoViD-19 epidemic comprises the period from February 26 (the first case) until April 3 (9 days after the partial quarantine introduction [5]). Phase 2 goes from March 24 (the partial quarantine introduction) to mid-June. Phase 3 lasts since mid-June (the relaxation introduction), and phase 4 could be initiated in phase 2 or 3 (the model adjusts the introduction time of more virulent variants).

Is the model capable of explaining the CoViD-19 epidemic in São Paulo State? Can the model predict the outcoming epidemic? We address these questions using equations (5) and (7) to estimate the model parameters against the CoViD-19 data collection from São Paulo State [9] (see Appendix B). From the accumulated cases and deaths estimated curves given by equations (6) and (8), we retrieve the daily cases and deaths using equations (9) and (10) and compare with the observed data.

### 3.1 Phases 1 and 2

To estimate the model parameters in phases 1 and 2, we use the data from February 26 to May 7 [5]. The estimated model parameters characterizing the natural epidemic and epidemic under quarantine are transported from Yang et al. [5]. Considering lower virulent pool of SARS-CoV-2 (*l*_*y*_ = *l*_*o*_ = 0.8), the values are:

1. **Transmission rates** (from February 26 to April 3) – Estimated values are *β*_*y*_ = 0.78 and *β*_*o*_ = 0.90 (both in *days*^−1^), resulting in the basic reproduction number *R*_0_ = 9.24. When the CoViD-19 epidemic was initiated, we did not have any control mechanisms, which is characterized as the natural epidemic. The effect of isolation on March 24 appears around 9 days later [5]. The initial conditions at *t* = 0 (February 26) supplied to the system of equations (1), and (3) are given by equation (A.3) in Appendix A

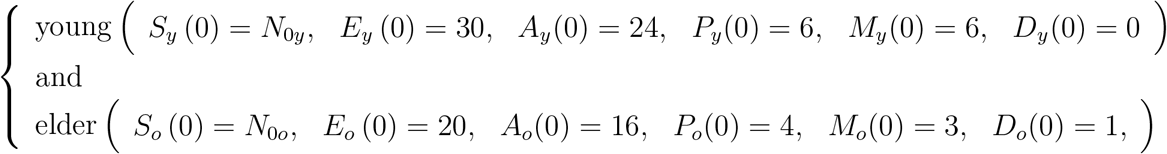

plus *Q*_*y*_ (0) = *Q*_*o*_ (0) = *R*(0) = 0. For São Paulo State, *N*_0*y*_ = 37.8 million and *N*_0*o*_ = 6.8 million. (See [5] for details in the initial conditions’ setup.)
2. **Proportion in partial quarantine** (from March 24 to April 13) – Estimated value is *u* = 0.53. The effect of adopting protective measures on April 4 appears around 9 days later. The boundary conditions at *t* = *τ* = 27 (March 24) supplied to the system of equations (1), and (3) are given by equation (A.4) in Appendix A,

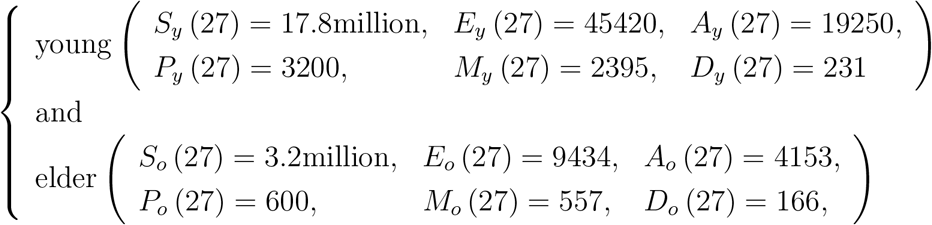

with *Q*_*y*_ (27) = 19.9 million, *Q*_*o*_ (27) = 3.59 million, and *R* = 10032. These conditions refer to the circulating subpopulation, and we assumed that only susceptible individuals are isolated.
3. **Protective measures** (from April 4 to May 7) – Estimated value is *ε* = 0.48. The circulating individuals adopted at *t* = 38 (April 4) the protective measures and social distancing, reducing the protective factor *ε* from one to 0.48.
4. **Additional mortality rates** (from March 16 to May 22) – Estimated values are *α*_*y*_ = 0.00185 and *α*_*o*_ = 0.0071 (both in *days*^−1^) fixing Δ = 15 *days*. The first death due to CoViD-19 was on March 16, and we considered 15 more observed data than previous estimations [5].

Step (1) estimates the natural epidemic curve *D*, providing the basic reproduction number *R*_0_. Steps (2) and (3) estimate the flattened epidemic curve by isolation (*u*) and protective measures by personal hygiene, use of face masks, and social distancing (*ε*). Finally, step (4) estimates the CoViD-19 fatality rates (*α*_*y*_ and *α*_*o*_).

Figure 1 shows the estimated curve Ω and the observed data Ω^*ob*^ (a), and the estimated curve Π and the observed data Π^*ob*^ (b) considering partial quarantine in São Paulo State. We stress that the curve Ω is the accumulated severe CoViD-19 cases, and Ω^*ob*^ is the accumulated CoViD-19 registered data encompassing all cases.

**Figure 1:**
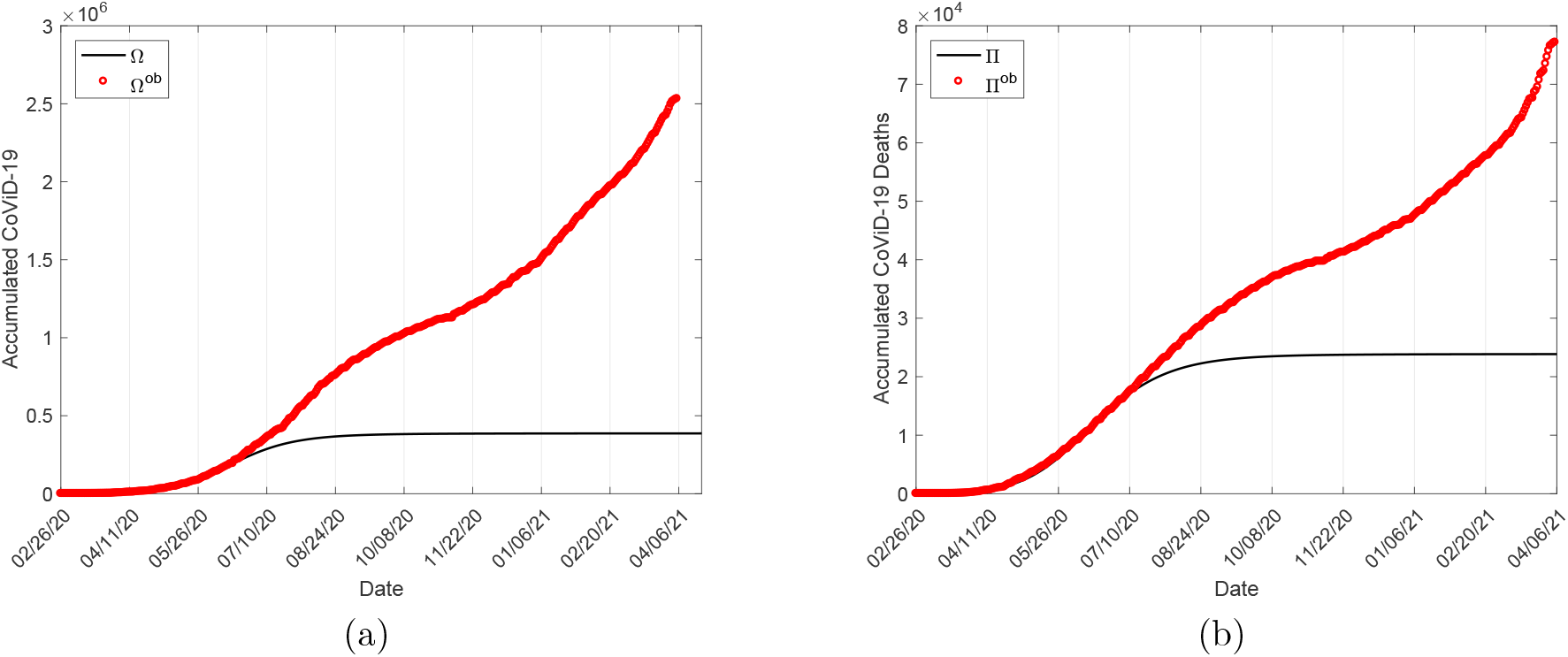
The estimated curve Ω and the observed data (a), and the estimated curve Π and the observed data (b) under partial quarantine in São Paulo State.

We address the questions of the capability to describe and forecast the CoViD-19 epidemic. The model parameters were estimated using collected data from February 26 to May 7 (cases) or May 22 (deaths). During this period, the model described the natural epidemic followed by the epidemic under isolation. Observing Figure 1, the estimated curves of Ω and Π fit the observed data until June 18 and July 9. In other words, the observed data were matched with the adjusted curve during the period from May 8 to June 18 for Ω (or May 23 to July 9 for Π). São Paulo State initiated but not fully implemented the relaxation in June 2020 (see the proportion in isolation in Figure B.3, Appendix B). Hence, the model showed the capability of predicting the outcoming epidemic while the isolation lasted – Figure 1 showed the registered data and the estimated Ω and Π curves separated when São Paulo State implemented the relaxation.

We assumed that among isolated individuals, SARS-CoV-2 is not transmitting. However, this is not what is occurring. The interaction between one person infected during the circulation and transmitting the virus to household and neighborhood people is very complex but can be incorporated into the model. This transmission increases as the number of individuals who circulate increases when the relaxation takes place. Instead of including influx and outflux of individuals from circulating to isolated subpopulations and vice-versa, we simplify the complex transmission between them by one input – from circulating to isolated subpopulations. For this reason, we let a low transmission among them some days before the epidemic’s peak, which occurred on June 27: On June 1, 2% of exposed and 7% of asymptomatic and pre-symptomatic individuals enter into isolated subpopulation.

Therefore, we consider SARS-CoV-2 transmission in two independent circulating and isolated subpopulations. The CoViD-19 epidemic in each subpopulation is driven by the system of equations (1), (2) and (3). The previously non-transmitting isolated population receives 2% of exposed and 7% of asymptomatic and pre-symptomatic individuals from the circulating subpopulation, triggering an epidemic. The initial conditions at *t* = 96 (June 1) in the isolated subpopulation are

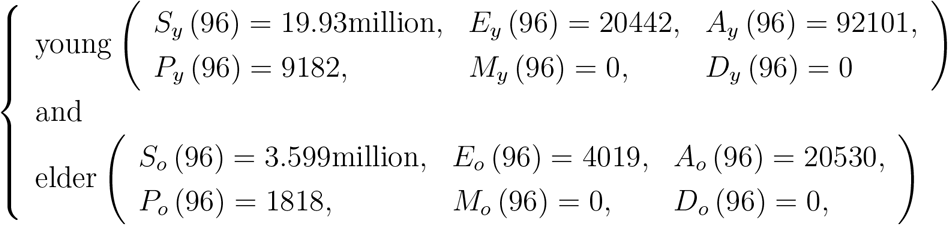

with *R* = 0. Observe that the compartment *Q* becomes susceptible individuals in the isolated subpopulation. At this time, the boundary conditions in the circulating subpopulation are

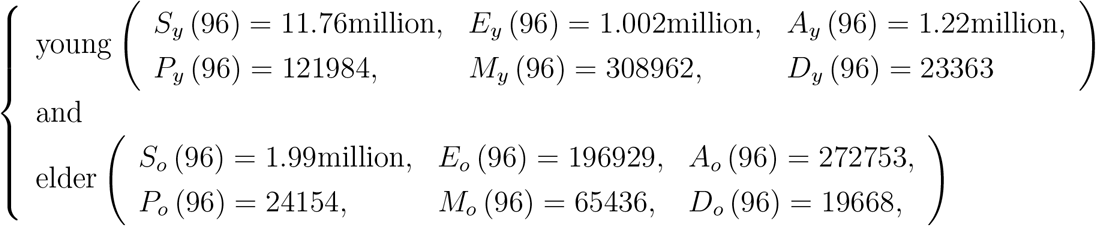

with *R* = 3.92 million. These conditions are the transfer of 2% of exposed and 7% of asymptomatic and pre-symptomatic individuals to isolated subpopulation. The initial and boundary conditions are similar than those equations (A.3) and (A.4) in Appendix A, but for variables *E, A* and *P*.

The new outbreak among isolated individuals is triggered considering the above estimated transmission rates, but reduced by the factor *ω*, see equation (A.1). Next, we consider the relaxation and how the mutation acts populationally.

### 3.2 Phases 3 and 4

Figure 1 summarized the epidemic under isolation. As pointed out, the absence of mass test allowed to estimate firstly the accumulated severe CoViD-19 curve Ω, from which the accumulated CoViD-19 deaths curve Π was estimated. The observed data were well-fitted to estimated Ω and Π curves. In June 2020, more tests were begun to caught mild and asymptomatic cases. The observed data Ω^*ob*^ also encompass mild and asymptomatic cases. Hence, we estimate the accumulated CoViD-19 deaths curve Π and retrieve the corresponding accumulated severe CoViD-19 curve Ω.

We assume that the isolated individuals’ release relaxes the protective measures, increasing the factor *ε* in the circulating subpopulation. In the isolated subpopulation, the relaxation increases the transmission among isolated individuals by diminishing the transmission factor *ω*. The reason is the close contact between circulating individuals when in family contact with isolated individuals. Besides relaxation, we explain the increased mortality considering mutations originating a pool with more virulent variants.

When a release occurs, isolated individuals are transferred to the circulating subpopulation. This release is described by the boundary conditions given by equation (A.5) in Appendix A. However, all compartments’ values are not changed when mutations occur, and more virulent variant circulates. In this case only the parameter related to the virulence *l*_*y*_ and *l*_*o*_ are varied [6].

As we have pointed out, the SARS-CoV-2 is transmitting independently in the circulating and isolated subpopulations before the beginning of the relaxation. Hence, the epidemic curve *D* is the sum of the severe CoViD-19 cases in the isolated 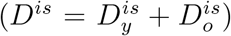 and circulating (*D* = *D*_*y*_ + *D*_*o*_) subpopulations.

We consider the relaxation initiating on June 30. At the moment of the release, we vary the protection factor *ε* in the circulating subpopulation and the reduction factor *ω* in the isolated subpopulation. If necessary, we also vary the fraction of asymptomatic individuals *l*. We consider 4 releases, but we present only the boundary conditions at the first release *t* = 125.

The circulating subpopulation receives 7% of individuals from isolated subpopulation; hence the boundary conditions are

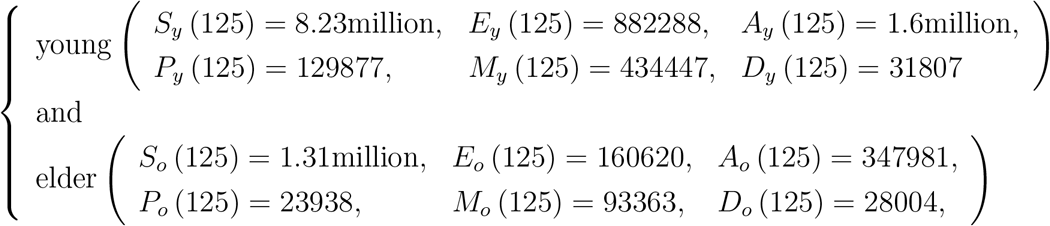

with *R* = 9.32 million. The isolated subpopulation is decreased in 7% of individuals, and the boundary conditions are

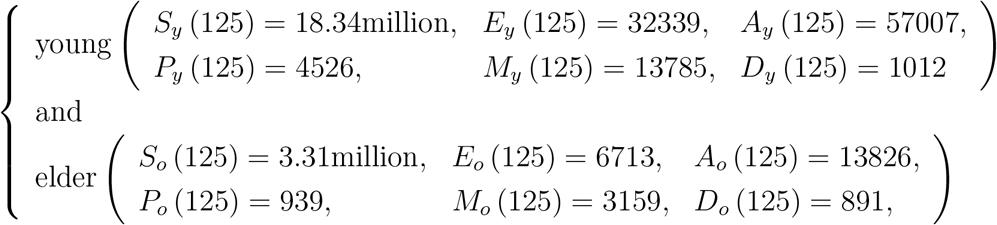

with *R* = 212230.

In phases 3 and 4, besides the relaxation, we consider the appearance of more virulent variant(s) of SARS-CoV-2. The main effect of a more virulent SARS-CoV-2 transmission is the increased number of fatalities due to severe CoViD-19. The combined effects of relaxation and virulence are evaluated by estimating the observed number of accumulated deaths, and the severe CoViD-19 curve *D* is retrieved from the estimated Π.

Table 1 shows the date of the *i*-th release, and the corresponding value for *u*_*i*_; and the variation in the protective measure *ε*, the decreased transmission factor among isolated individuals *ω*, and the fraction of asymptomatic individuals *l* and the corresponding dates to estimate the observed CoViD-19 deaths. At the time of release, the parameters *ε, ω*, and *l* are varied together. However, further variations in *ε, ω*, and *l* are needed to adjust the observed deaths Π^*ob*^.

**Table 1:** Interventions since the beginning of the relaxation (*u*_*i*_). In the circulating subpopulation, the protective measues is relaxed(*ε*), and in the isolated subpopulation, the transmission increases (*ω*). In both subpopulations, the virulence increases (*l*).

| Date | $u_i$ | $\varepsilon$ | $\omega$ | $l$ |
| --- | --- | --- | --- | --- |
| During isolation | 0 | 0.48 | — | 0.8 |
| June 1, 2020 | 0 | 0.48 | 10 | 0.8 |
| June 30, 2020 | 0.07 | 0.6 | 10 | 0.78 |
| August 4, 2020 | 0.07 | 0.7 | 9 | 0.65 |
| August 24, 2020 | 0.05 | 0.75 | 8 | 0.55 |
| September 13, 2020 | 0.05 | 0.75 | 7 | 0.45 |
| October 3, 2020 | 0 | 0.75 | 6.5 | 0.4 |
| October 23, 2020 | 0 | 0.75 | 5.8 | 0.35 |
| January 6, 2021 | 0 | 0.75 | 5 | 0.35 |
| March 2, 2021 | 0 | 0.75 | 4 | 0.25 |

Based on the values given in Table 1, Figure 2 shows the estimated curve Π and the observed data Π^*ob*^ (a), and the retrieved accumulated severe CoViD-19 curve Ω and the observed data Ω^*ob*^ (b). The release of the isolated people, the transmission in the isolated subpopulation, and the occurrence of mutations fitted the observed data since the relaxation. Figure 2 is the complement of the fitting shown in Figure 1.

**Figure 2:**
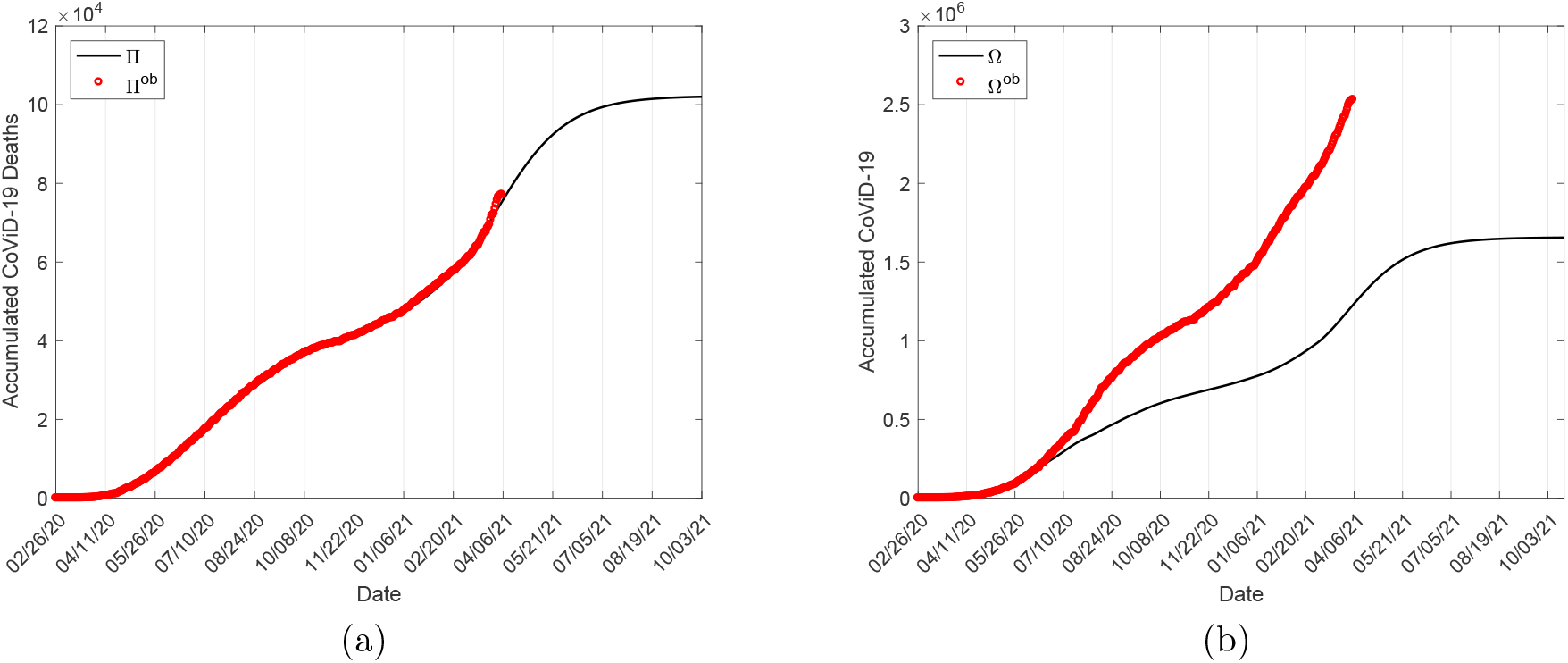
The estimated curve Π and the observed data (a), and the estimated curve Ω and the observed data (b).

The estimated curve Π fits very well the observed data (Figure 2(a)), but the retrieved curve Ω (Figure 2(b)) from estimated curve Π situates below the observed CoViD-19 cases. The observed data Ω^*ob*^ contain all SARS-CoV-2 infections caught by tests, but the curve Ω is related to the severe CoViD-19 cases. These estimated curves Π and Ω explained the CoViD-19 epidemic under isolation, relaxation, and appearance of virulent variants from February 26, 2020, to April 5, 2021. To assess the model’s predictability, the curves Π and Ω were extended until they approach asymptotes (October 2021). At the end of the epidemic, the asymptotic values are 102000 and 1.66 million for deaths and severe CoViD-19, resulting in the severity cases fatality rate of 6.15%. The accumulated new infection cases at the plateaux are 36.64 million, resulting in an infection fatality rate of 0.28%.

Figure 2 shows two sequential sigmoid-shaped Π and Ω curves. In the sigmoid curve, the upward concavity is changed to downward concavity at the inflection point (time) [10] before reaching the plateaux. This behavior corresponds to the variation in the effective reproduction number *R*_*ef*_ – In the upward concavity, we have *R*_*ef*_ *>* 1, *R*_*ef*_ = 1 at the inflection time, and *R*_*ef*_ *<* 1 in the downward concavity. In other words, the upward concavity corresponds to the ascending phase of the daily cases Ω_*d*_, and the downward concavity to the falling phase. The São Paulo State CoViD-19 epidemic presents two sequential sigmoid curves, with the transition from the first to the second sigmoid occurring around November 10, 2020 (Figure 2(a)). At this transition time, when *R*_*ef*_ *<* 1 changes to *R*_*ef*_ *>* 1, the first sigmoid was approaching the plateaux.

A second sigmoid’s appearance reveals turn-over in the epidemic, which is analyzed assuming everlasting immunity. The second sigmoid initiating around November 10 showed a slower followed by a rapid increase than the first sigmoid (Figure 2(b)). Hence, this “new outbreak” can not be explained by relaxation alone. The incorporation of virulence and transmission in the isolated subpopulation successfully explained the epidemic in São Paulo State. Summarizing:

1. **The first sigmoid curve**, from February 26 to November 10, 2020 – This curve characterizes the partial quarantine and relaxation.
2. **The second sigmoid curve**, from November 11, 2020, to April 5, 2021 – This curve characterizes the relaxation, the transmission in the isolated subpopulation, and the transmission of more virulent variants.

### 3.3 Quarantine, relaxation, and mutation

Figure 2 portrayed the CoViD-19 epidemic in São Paulo State from February 26, 2020, to April 5, 2021. In this period, São Paulo State implemented partial quarantine, and a timid relaxation began, but not fully implemented, and isolation persists up to now. To better understand this epidemiological scenario, we retrieve more curves related to the CoViD-19 epidemic from the estimated Π and Ω curves.

Figure 3 shows the daily deaths curve Π_*d*_ and the observed data 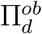 (a), and the daily CoViD-19 cases curve Ω_*d*_ and the observed data 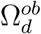 (b). The daily cases and deaths are calculated using equations (10) and (9). We also show the 7-day moving average, which is close to the estimated curve. The outliers in the 7-day moving average fatalities observed at the end of March and beginning of April 2021 can be understood as the surpassed deaths due to the lack of hospital care due to the huge number of severe CoViD-19 cases. Another possible explanation is the transmission of a more virulent variant.

**Figure 3:**
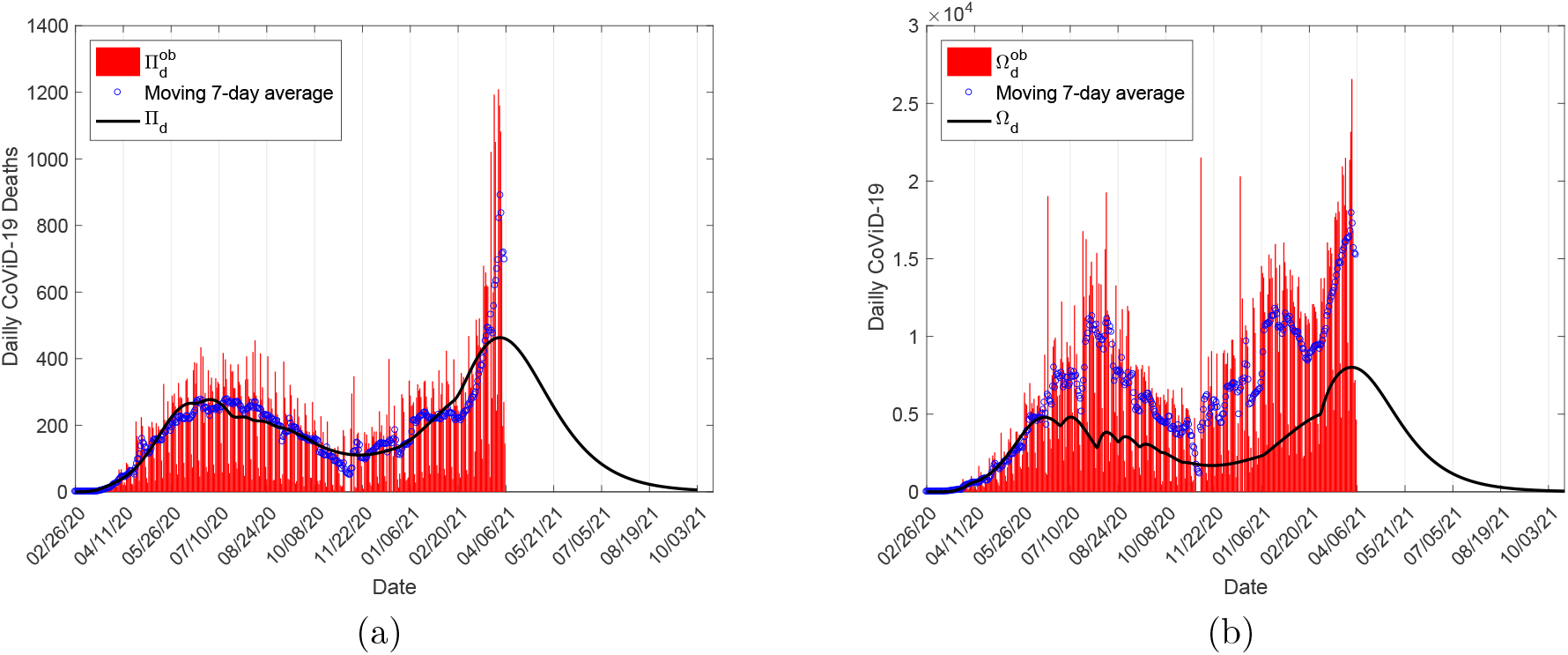
The daily deaths curve Π_*d*_ and the observed data (a), and the daily cases curve Ω and the observed data (b). We also show the 7-day moving average.

In the first epidemic, the peaks of Π_*d*_ and Ω_*d*_ are 277 and 4808, occurring on July 2 and July 1, 2020. This epidemic corresponds to the first sigmoid shown in Figure 2 and characterizes partial quarantine and part of the relaxation. In the epidemic corresponding to the second sigmoid, the peaks of Π_*d*_ and Ω_*d*_ are 463 and 8015, occurring on April 1, 2021. This epidemic characterizes the relaxation, transmission in isolated individuals, and the transmission of more virulent variants of SARS-CoV-2.

The role played by the transmission in the isolated subpopulation can be seen in the epidemic curve. Figure 4 shows the epidemic curve *D* for young, elder, and total population in the circulating (continuous line) and isolated (dashed line) subpopulations (a), and the entire population (b).

**Figure 4:**
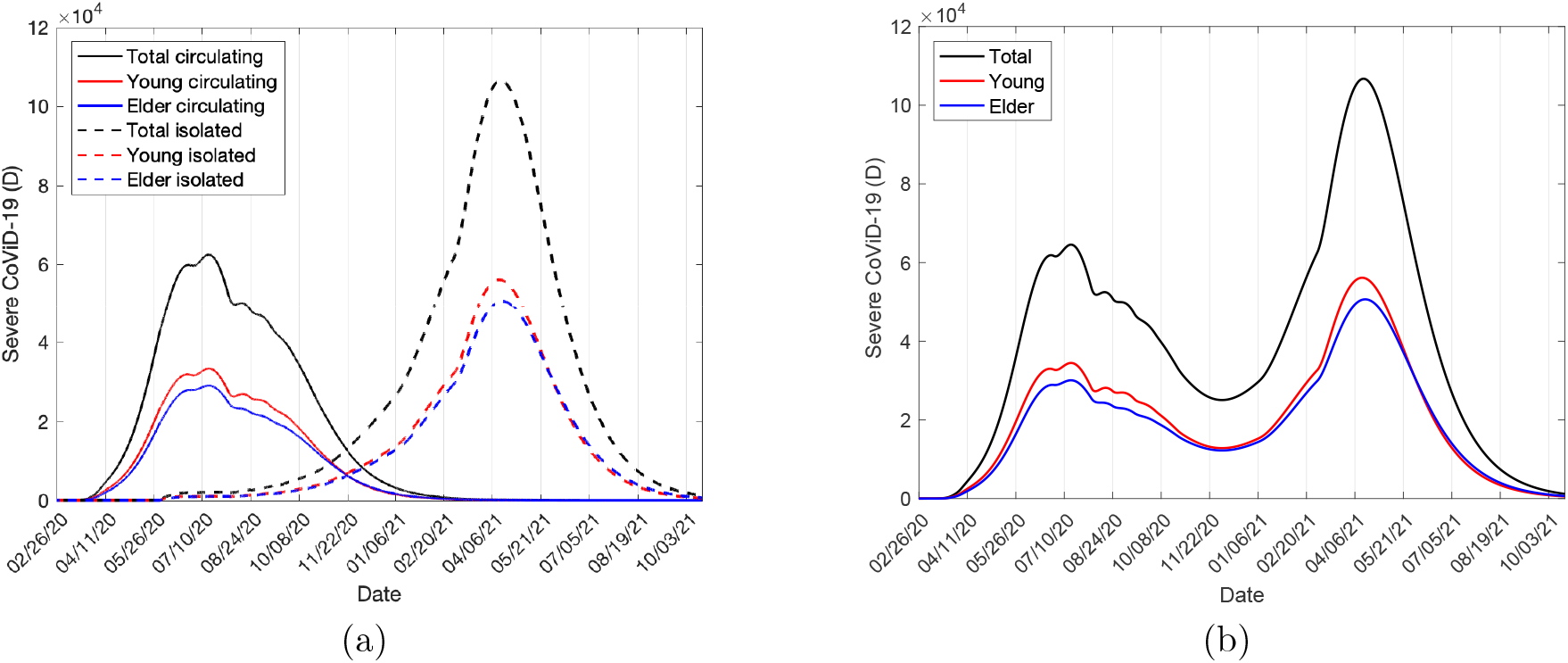
The epidemic curve *D* for young, elder and total population in the circulating (continuous line) and isolated (dashed line) subpopulations (a), and the entire population (b).

The peaks of *D* corresponding to the first and second sigmoid curves are 64550 and 106700, occurring on July 16, 2020, and April 14, 2021. The population in isolation was 47%, but the originally circulating 53% of susceptible individuals were reduced by infection (see Figure 4(a)). Hence, the increase in 65% of the severe CoViD-19 cases can not be explained by relaxation alone. On the contrary, the majority of the infections occur due to transmission in the isolated subpopulation. Hence, the transmission in the isolated subpopulation and the enhanced virulence resulted in the increased fatalities during the second sigmoid epidemic phase (see Figure 3).

Figure 5 shows the susceptible individuals *S* for young, elder, and total population in the circulating (continuous line) and isolated (dashed line) subpopulations (a), and the entire population (b).

**Figure 5:**
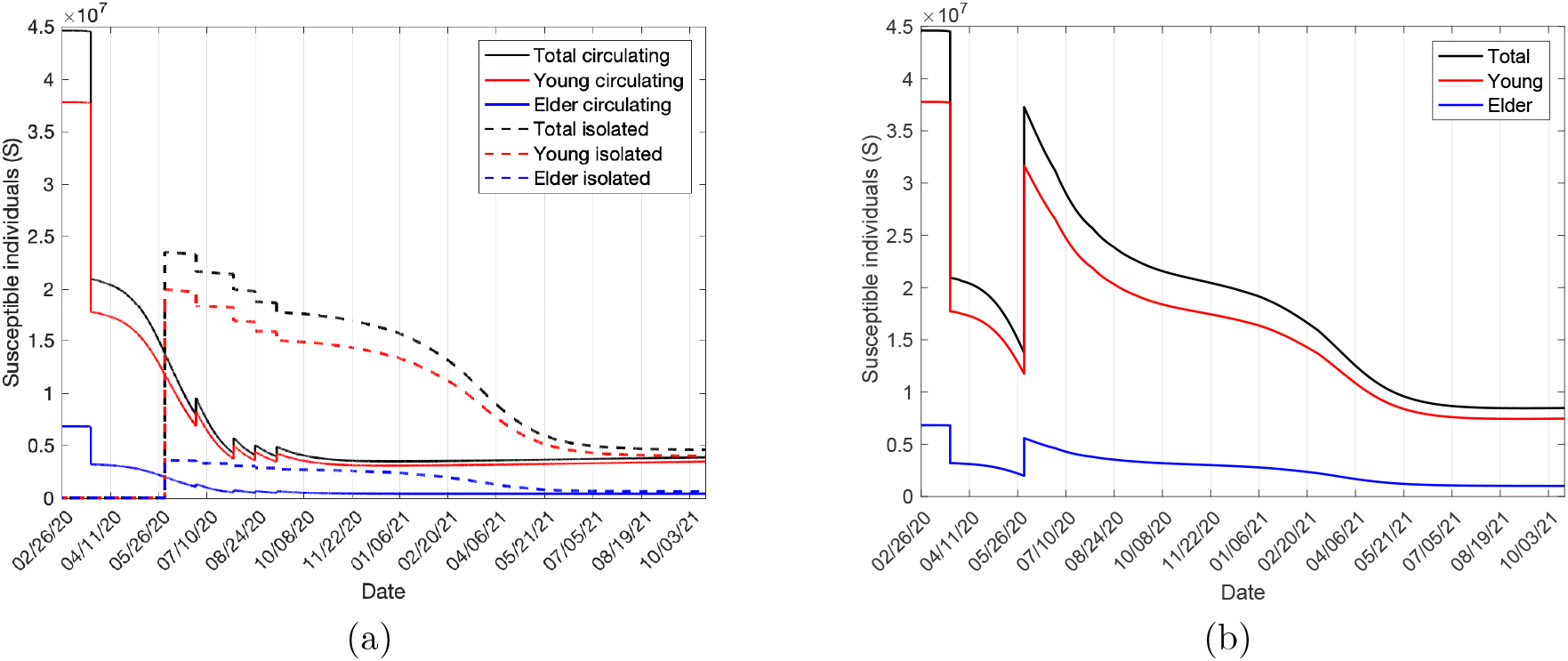
The susceptible individuals *S* for young, elder and total population in the circulating (continuous line) and isolated (dashed line) subpopulations (a), and the entire population (b).

When the epidemic approaches the plateaux, the susceptible individuals *S* for the circulating and isolated subpopulations are 3.87 million and 4.60 million. The reminiscent susceptible population is 19% of the population, indicating a potential retaken but a weakened epidemic.

Figure 6 shows the immune individuals *R* for young, elder, and total population in the circulating (continuous line) and isolated (dashed line) subpopulations (a), and the entire population (b).

**Figure 6:**
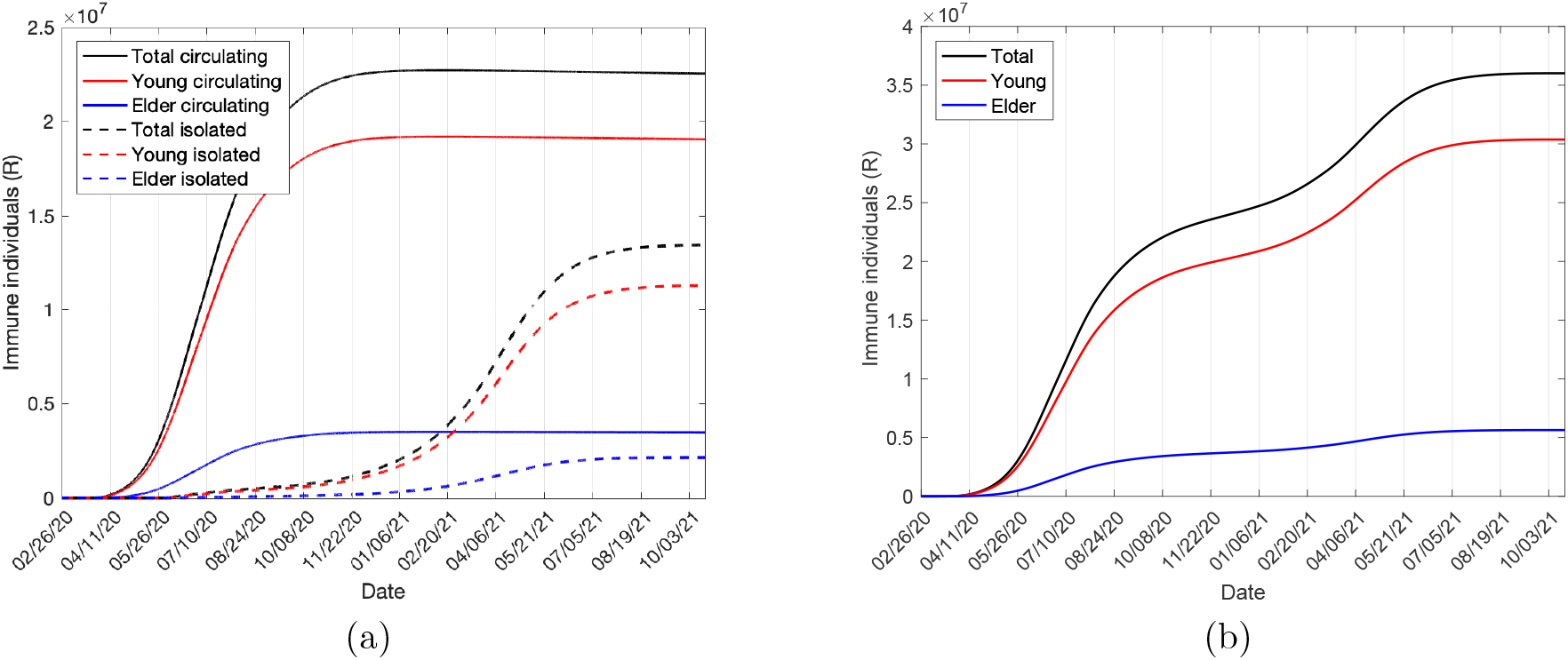
The immune individuals *R* for young, elder and total population in the circulating (continuous line) and isolated (dashed line) subpopulations (a), and the entire population (b).

When the epidemic approaches the plateaux, the immune individuals *R* for the circulating and isolated subpopulations are 22.55 million and 13.46 million. In comparison with the natural epidemic’s immune individuals (97% of the population [5]), 81% of the immune individuals show that around 16% of the population are at risk if all protective measures could be abandoned.

The model given by equations (1), (2) and (3) described the epidemiological scenario in São Paulo State from February 26, 2020 to April 5, 2021 (see Figure 2(a)). On April 5, the susceptible individuals *S* for the circulating and isolated subpopulations are 3.58 million and 9.01 million (28% of the population), and the immune individuals *R* for the circulating and isolated subpopulations are 22.71 million and 7.18 million (67% of the population). Additionally, the number of deaths is 75700, which is 98.1% of 77165 observed fatalities. The epidemiological curves presented in Figures 3-6 showed that the CoViD-19 epidemic in São Paulo State reached the peak. Using the estimated parameters, the model is tested in its capacity to predict an outcoming epidemic. For this purpose, we extended the estimated curves from April 6 to October 2021 (see Figure 2). The extended epidemiological curves presented in Figures 3-6 show that the CoViD-19 epidemic in São Paulo State initiated the descending phase.

Is the virulence essential to explain the CoViD-19 epidemic in São Paulo State? Figure 7 shows the release scheme presented in Table 1, except the appearance of more virulent variants – The estimated curve Π and the observed data (a), and the retrieved curve of accumulated severe CoViD-19 cases Ω and the observed data (b) without change in the virulence. Hence, during the entire epidemic, the virulence was fixed in *l* = 0.8.

**Figure 7:**
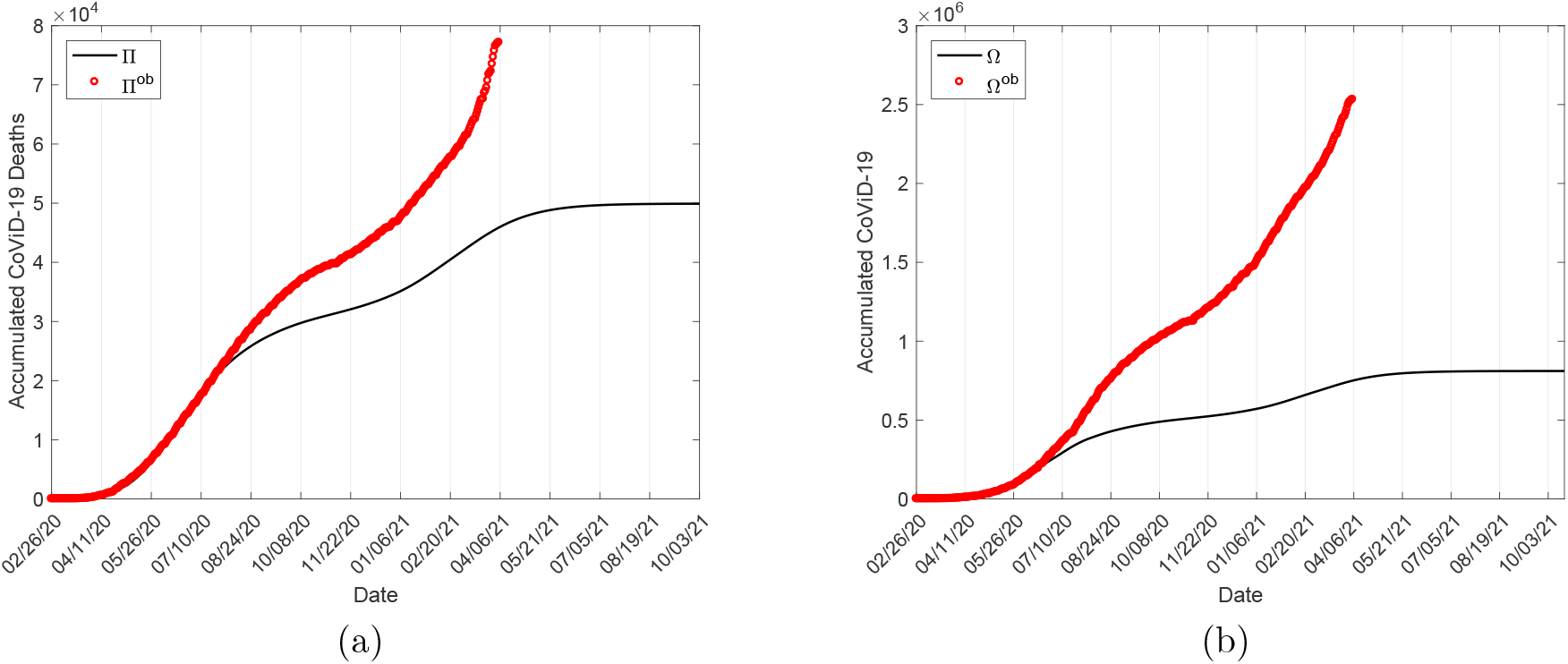
The estimated curve Π and the observed data (a), and the estimated curve Ω and the observed data (b).

Suppose we neglect the appearance of a more virulent variant. At the end of all individuals’ release, the susceptible and immune individuals approach 16.1% and 83.8% of the population, and the number of fatalities, 49910. Hence, model-based analysis of the data showed the necessity of increased deaths probably due to enhanced virulence.

## 4 Discussions

In all preceding results, we considered parameters presented in Table A.2 in Appendix A and the estimated additional mortality rates using the fatality data from March 16 to May 22, 2020, unchanging in the epidemic. However, some of these parameters can be changed as we allowed parameters in Table 1 to vary.

The mutations occur during the replication, which can be incorporated in the model, letting a fraction of infected individuals producing mutations during the virus replication. However, we simplified by assuming that the ratio between asymptomatic and symptomatic individuals (parameter *l*) mimics the level of virulence: The larger the proportion of symptomatic, the higher is the virulence. The epidemiological scenario in Amazonas State (Brazil) can shed more light on the virulent variants’ role in association with relaxation. In Amazonas State, the occurrence of deaths in January-February, 2021, is higher than that occurred during all 2020 [11]. This increased number can not be explained only by the partial relaxation once a significant number of deaths caused by the P.1 variant was found.

The complex and hardly measurable interaction between individuals in circulation and his/her family and neighbor in isolation were oversimplified, assuming that a new epidemic was triggered at a specific time. The second sigmoid’s upward concavity presented a higher increase than the first sigmoid phase, showing that the relaxation can not explain the observed data, even we relaxed the protective measures (*ε*). Hence, the transmission among isolated individuals (parameter *ω*) is crucial to explain the second sigmoid curve observed in the accumulated fatalities due to CoViD-19.

We analyzed only the case where the same proportion of young and elder subpopulations is isolated and released (parameters *u* and *u*_*i*_). Young individuals are circulating due to their work more than elders. Then, the isolation in the young subpopulation is lower, but the release is higher than in the elder subpopulation. This characteristic can be incorporated into the model.

The model did not consider the loss of immunity, so the transmission among isolated subpopulation and the enhanced virulence explained the increased mortality. The waning of immunity can be incorporated in the model to assess the role of reinfection [7]. Also, the impact of vaccination on the epidemic can be evaluated by the model. In March 2021, São Paulo State initiated the vaccination of health workers and the elder subpopulation.

The estimation of the model parameters using the accumulated data showed superior to the estimation based on the daily recorded CoViD-19 data [12]. Figures B.1(a) and B.2(a) showed irregular recording patterns, especially during the weekends. In contrast, the accumulated CoViD-19 data (Figures B.1(b) and B.2(b)) showed smoothed curves. Notably, the daily CoViD-19 cases obtained from the estimated Ω curve and the 7-day moving average matched reasonably well, according to Figure 3(a). Moreover, the estimated Ω and Π curves showed the epidemic’s main characteristic – The sigmoid-shaped curve. This pattern permitted us to observe a second sigmoid curve and analyze the model according to the properties carried on by this curve – The quick increase in the epidemic is associated with a higher risk of infection, which is associated with a higher *R*_0_ [10]. From Figure 2, the epidemic in the second sigmoid increased faster than that observed in the first sigmoid. Hence, one explanation for the São Paulo State’s epidemiological scenario could be a higher *R*_0_ for the CoViD-19 epidemic and the transmission among isolated individuals.

Sabino et al. [7] provided another approach to describe the increased CoViD-19 fatalities. To explain the CoViD-19 epidemic in Amazonas State, they stated in one of the hypotheses that “the SARS-CoV-2 attack rate could have been overestimated during the first wave, and the population remained below the herd immunity threshold until the beginning of December 2020”. They concluded by assuming *R*_0_ = 3 and, consequently, the herd immunity is 1 − 1*/R*_0_ = 0.67. The herd immunity concept (the percentage of susceptible individuals being immunized by vaccine or removed from the infection chain to eliminate the transmission) is valid only when the epidemic reaches a steady-state. However, this concept can be applied in the CoViD-19 epidemic’s first wave: How many susceptible individuals must be isolated to break the transmission. For *R*_0_ *>* 3, the transmission fades out after almost all individuals being infected (by simulating the model, recovered individuals are 95% of the population); hence the “herd immunity” is achieved when 5% of susceptible individuals are at risk. Therefore, higher *R*_0_ associated with increased virulence and transmission among isolated individuals may better describe the CoViD-19 epidemic in Amazonas State, as we did for São Paulo State.

Although the model proposed here was an over-simplified description of the highly complex disease propagation phenomenon, we obtained a good explanation of the CoViD-19 epidemic in São Paulo State. This goal was attained by estimating the model parameters related to the partial quarantine, protective measures, transmission among isolated people, and virulent variants’ appearance during the epidemic. The model also showed predicting characteristics. Summarizing:

1. The model provided a well-description of the epidemic under isolation and predicted the outcoming epidemic while the partial quarantine lasted. The epidemic under isolation was well-described by a lower virulent SARS-CoV-2 transmission.
2. The further epidemic when the relaxation initiated, the epidemic was better described with the transmission in the isolated subpopulation and enhanced virulence until April 5, 2021. The model predicted that the epidemic was in the descending phase, but another 26300 fatalities are expected at its end if the mass vaccine will not occur.

## 5 Conclusion

We developed a deterministic model, given by equations (1), (2) and (3), to describe the SARS-CoV-2 transmission. The model was formulated to assess the impacts of the quarantine, relaxation, and virulence on the CoViD-19 epidemic. This model was applied to explain the CoViD-19 epidemic in São Paulo State. Notwithstanding its simplicity, the model fitted the CoViD-19 data and permitted the outcoming epidemic’s prediction.

The scenario characterizing the São Paulo State CoViD-19 epidemic presented here was obtained by a deterministic model. This simplified model can be improved in several ways to provided more reliable epidemiological scenarios. The change in the CoViD-19 natural history’s parameters according to the epidemiological status. The inclusion of waning immunity and the vaccination in the model. Introduction of more realistic descriptions of the mutations and the interaction between circulating and isolated individuals in the model. These improvements are left to further works.

## Data Availability

Data available at Health ministry site

https://covid.saude.gov.br/

## Availability of data and material

The data that support the findings of this study are openly available in SP contra o novo coronavírus (Boletim completo) at https://www.seade.gov.br/coronavirus/.

## Author contributions

Hyun Mo Yang: Conceptualization, Methodology, Formal analysis, Writing - Original draft preparation, Validation, Supervision. Luis Pedro Lombardi Junior: Software, Data Curation, Visualization, Validation. Ariana Campos Yang: Conceptualization, Validation, Investigation.

## A The SQEAPMDR model

One of the main aspects of CoViD-19 is increased fatality in the elder subpopulation. For this reason, a population is divided into two groups, composed of young (60 years old or less, denoted by subscript *y*) and elder (60 years old or more, denoted by subscript *o*) subpopulations. This community’s vital dynamic is described by the per-capita rates of birth (*ϕ*) and mortality (*µ*), and *φ* is the aging rate, that is, the flow from young subpopulation *y* to elder subpopulation *o*. Another aspect is the presence of the pre-symptomatic individuals, that is, individuals without symptoms transmitting SARS-CoV-2 before the onset of the disease [13].

Hence, for each subpopulation *j* (*j* = *y, o*), individuals are divided into seven compartments: susceptible *S*_*j*_, isolated (quarantine) *Q*_*j*_, exposed and incubating *E*_*j*_, asymptomatic *A*_*j*_, pre-symptomatic (or pre-diseased) individuals *P*_*j*_, symptomatic individuals with mild CoViD-19 *M*_*j*_, and severe CoViD-19 *D*_*j*_. However, all young and elder individuals in classes *A*_*j*_, *M*_*j*_, and *D*_*j*_ enter into the same recovered class *R* (this is the 8^*th*^ class, but common to both subpopulations). Table 1 summarizes the model compartments (dynamic variables).

The natural history of CoViD-19 is the same for young (*j* = *y*) and elder (*j* = *o*) subpopulations. We assume that individuals in the asymptomatic (*A*_*j*_), pre-diseased (*P*_*j*_), and a fraction *z*_*j*_ of mild CoViD-19 (*M*_*j*_) classes are transmitting the virus. Other infected classes ((1 − *z*_*j*_) *M*_*j*_ and *D*_*j*_) are under voluntary or forced isolation. Susceptible individuals are infected at a rate λ_*j*_*S*_*j*_ (known as the mass action law [14]), where λ_*j*_ is the per-capita incidence rate (or force of infection) defined by λ_*j*_ = λ (*δ*_*jy*_ + *ψδ*_*jo*_), with λ being

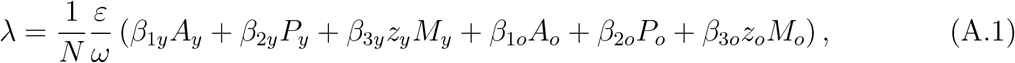

**Table A.1:**
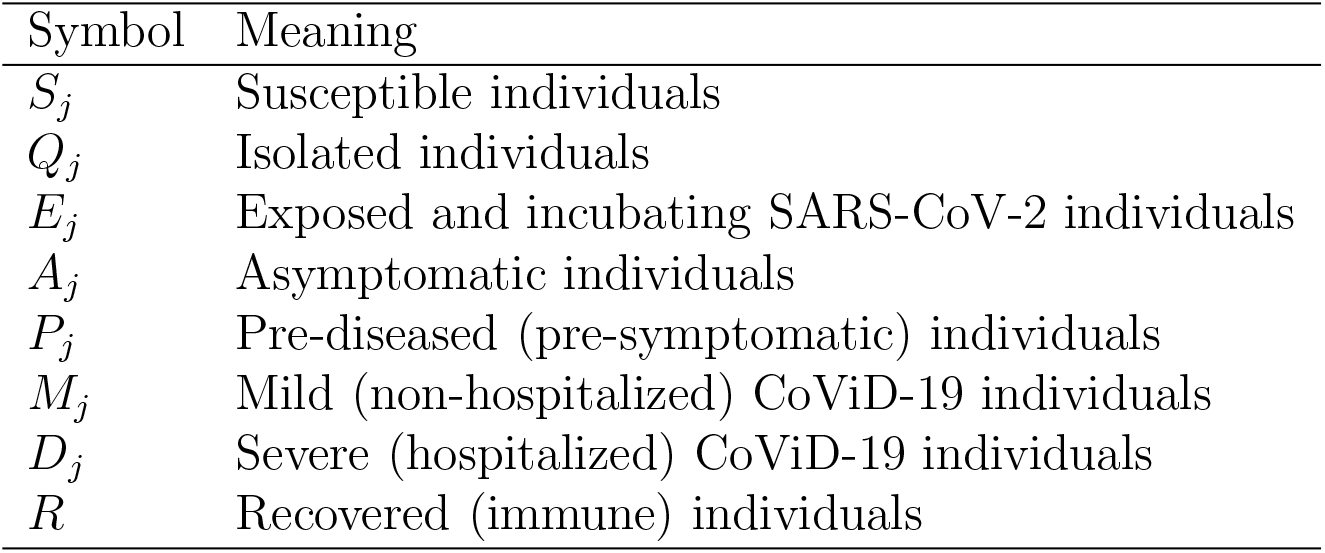
Summary of the model variables, for *j* = *y* (young) and *j* = *o* (elder).

where *δ*_*ij*_ is the Kronecker delta, with *δ*_*ij*_ = 1 if *i* = *j*, and 0, if *i* ≠ *j*; and *β*_1*j*_, *β*_2j_ and *β*_3*j*_ are the transmission rates, that is, the rates at which a virus encounters a susceptible people and infects him/her. The parameters *ε* ≤ 1 and *ω* ≥ 1 diminish the transmission rates – The protection factor *ε* decreases the transmission of infection by individual (face mask, hygiene, etc.) and collective (social distancing) protective measures, while the reduction factor *ω* decreases the transmission by the contact being restricted in the population in isolation (household and neighborhood contacts). In [3], a particular model was analyzed letting *z*_*y*_ = *z*_*o*_ = 0 and *χ*_*y*_ = *χ*_*o*_ = 1.

Susceptible individuals are infected at a rate λ_*j*_ and enter into class *E*_*j*_. After an average period 1*/σ*_*j*_ in class *E*_*j*_, where *σ*_*j*_ is the incubation rate, exposed individuals enter into the asymptomatic class *A*_*j*_ (with probability *l*_*j*_) or pre-diseased class *P*_*j*_ (with probability 1 − *l*_*j*_). After an average period 1*/γ*_*j*_ in class *A*_*j*_, where *γ*_*j*_ is the recovery rate of asymptomatic individuals, asymptomatic individuals acquire immunity (recovered) and enter into recovered class *R*. Possibly asymptomatic individuals can manifest symptoms at the end of this period, and a fraction 1 − *χ*_*j*_ enters into mild CoViD-19 class *M*_*j*_. For symptomatic individuals, after an average period 1*/γ*_1*j*_ in class *P*_*j*_, where *γ*_1*j*_ is the infection rate of pre-diseased individuals, pre-diseased individuals enter into severe CoViD-19 class *D*_*j*_ (with probability 1 − *k*_*j*_) or mild CoViD-19 class *M*_*j*_ (with probability *k*_*j*_). Individuals in class *D*_*j*_ acquire immunity after a period 1*/γ*_2*j*_, where *γ*_2*j*_ is the recovery rate of severe CoViD-19, and enter into recovered class *R* or die under the disease-induced (additional) mortality rate *α*_*j*_. Individuals in mild CoViD-19 class *M*_*j*_ acquire immunity after a period 1*/γ*_3*j*_, where *γ*_3*j*_ is the recovery rate of mild CoViD-19, and enter into recovered class *R*.

The dynamic equations for the compartments harboring virus *E, A, P, M*, and *D*, and recovered *R* are obtained through the balance between inflow and outflow in each compartment (see equations (2) and (3) in the main text). We assume that a unique pulse in isolation is adopted at time *t* = *τ*, described by *u*_*j*_*S*_*j*_*δ* (*t* − *τ*), and a series of intermittent releases, described by ∑_i=1_ *u*_*ij*_*Q*_*j*_*δ* (*t* − *t*_*ij*_), where 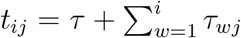 for *j* = *y, o*. The Dirac delta function *δ* (*x*) is *δ* (*x*) = ∞, if *x* = 0; otherwise, *δ* (*x*) = 0, with 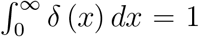. The fraction of persons in isolation is *u*_*j*_, and *u*_*ij*_, *i* = 1, 2, …, is the fraction of the *i*-th release of isolated persons, with *τ*_*wj*_ being the time between successive releases. The subsequent relaxation is described by a series of pulses. The isolation and further relaxation are translated as

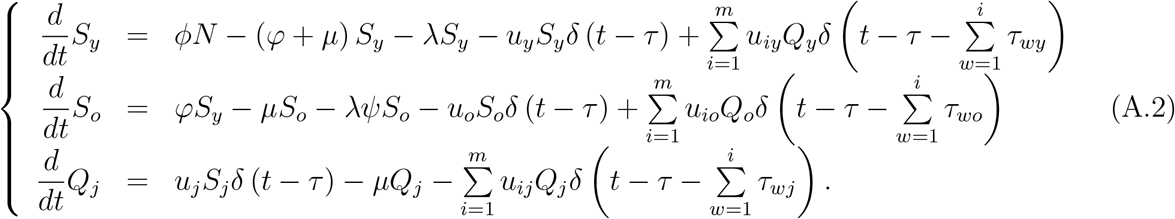

Table 2 summarizes the model parameters. The description of the assigned values to the CoViD-19 natural history parameters can be found in [5]. The values of the parameters not shown in the table are estimated considering two SARS-CoV-2 virulence levels.

The pulses in system of equations (A.2) are simulated permitting intermittent interventions to the boundary conditions. We drop out the Dirac delta functions, and we simulate the system of equations (1), (2), and (3) using the following initial and boundary conditions.

The initial conditions (simulation time *t* = 0) supplied to the system of equations (1), (2), and (3) are, for *j* = *y, o*,

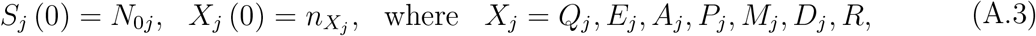

where 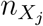 is a non-negative number. For instance, 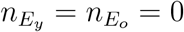 means that there is not any exposed person (young and elder) at the beginning of the epidemic. The isolation implemented at *τ* = 27 (corresponding to calendar time March 24, 2020) is described by the boundary conditions

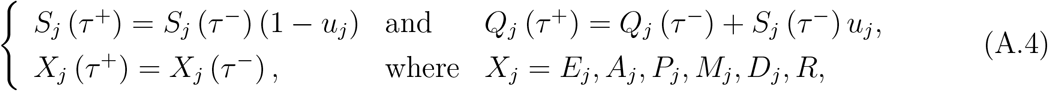

with *τ* ^−^ = lim_*t*→*τ*_ *t* (for *t < τ*), and *τ* ^+^ = lim_*τ*←*t*_ *t* (for *t > τ*). Each pulse of releases applied at a successive interval of time *τ*_*i*_, for *i* = 1, 2, …, is described by the boundary conditions supplied to the system at 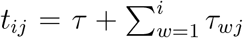. Hence, the boundary conditions for a series of pulses of release at *t*_*i*_, for *i* = 1, 2, …, *m*, are

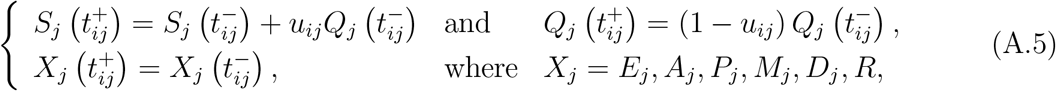

**Table A.2:** Summary of the model parameters (*j* = *y, o*) and values (rates in *days*^−1^ and proportions are dimensionless). (*) Values fitted considering the SARS-CoV-2 data from Saõ Paulo State.

| Symbol | Meaning | Value |
| --- | --- | --- |
| $\mu$ | Natural mortality rate | $1/(78.4 \times 365)$ |
| $\phi$ | Birth rate | $1/(78.4 \times 365)$ |
| $\varphi$ | Aging rate | $6.7 \times 10^{-6}$ |
| $\sigma_y (\sigma_o)$ | Incubation rate | $1/5.8 (1/5.8)$ |
| $\gamma_y (\gamma_o)$ | Recovery rate of asymptomatic individuals | $1/12 (1/14)$ |
| $\gamma_{1y} (\gamma_{1o})$ | Infection rate of pre-symptomatic individuals | $1/4 (1/4)$ |
| $\gamma_{2y} (\gamma_{2o})$ | Recovery rate of severe CoViD-19 individuals | $1/12 (1/21)$ |
| $\gamma_{3y} (\gamma_{3o})$ | Infection rate of mild CoViD-19 individuals | $1/13 (1/16)$ |
| $\tau$ | Time of the introduction of isolation | March 24, 2020 |
| $\tau_i$ | Period between $i$-th successive release | * |
| $z_y (z_o)$ | Proportion of transmission by mild CoViD-19 individuals | 0.5 (0.2) |
| $\chi_y (\chi_o)$ | Proportion of remaining as asymptomatic individuals | 0.98 (0.95) |
| $l_y (l_o)$ | Proportion of asymptomatic individuals | * |
| $k_y (k_o)$ | Proportion of mild (non-hospitalized) CoViD-19 | 0.92 (0.75) |
| $\varepsilon$ | Protection factor by protective measures | * |
| $\omega$ | Reduction factor by isolation | * |
| $u_y (u_o)$ | Proportion in isolation | * |
| $u_{iy} (u_{io})$ | Proportion released at $i$-th relaxation | * |
| $\alpha_y (\alpha_o)$ | Additional mortality rate | * |
| $\beta_{1y} (\beta_{1o})$ | Transmission rate due to asymptomatic individuals | * |
| $\beta_{2y} (\beta_{2o})$ | Transmission rate due to pre-symptomatic individuals | * |
| $\beta_{3y} (\beta_{3o})$ | Transmission rate due to mild CoViD-19 individuals | * |

where we have 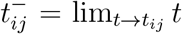, and 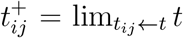 (for *t* > *t*_*ij*_). If *τ*_*wj*_ = *τ*_*j*_, then *t*_*ij*_ = *τ* + *iτ*_*j*_.

The system of equations (1), (2) and (3) does not reach steady-state (time varying population). However, this system in term of fractions attains steady-state. Using the next generation matrix theory to the trivial equilibrium point *P* ^0^, and applying the method proposed in [15] and proved in [16], we obtained the basic reproduction number *R*_0_ given by

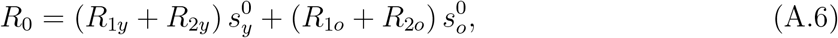

where the partial basic reproduction numbers *R*_1*y*_, *R*_2*y*_, *R*_1*o*_, and *R*_2*o*_ are given by equation (12) in the main text, and the initial fractions are 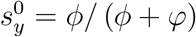 and 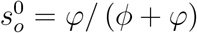. Details can be found in [5].

## B CoViD-19 data and isolation

We present the data collected from São Paulo State [9].

Figure B.1 shows the daily (a) and accumulated (b) CoViD-19 records, including severe cases and those (mild and asymptomatic cases) caught by test.

**Figure B.1:**
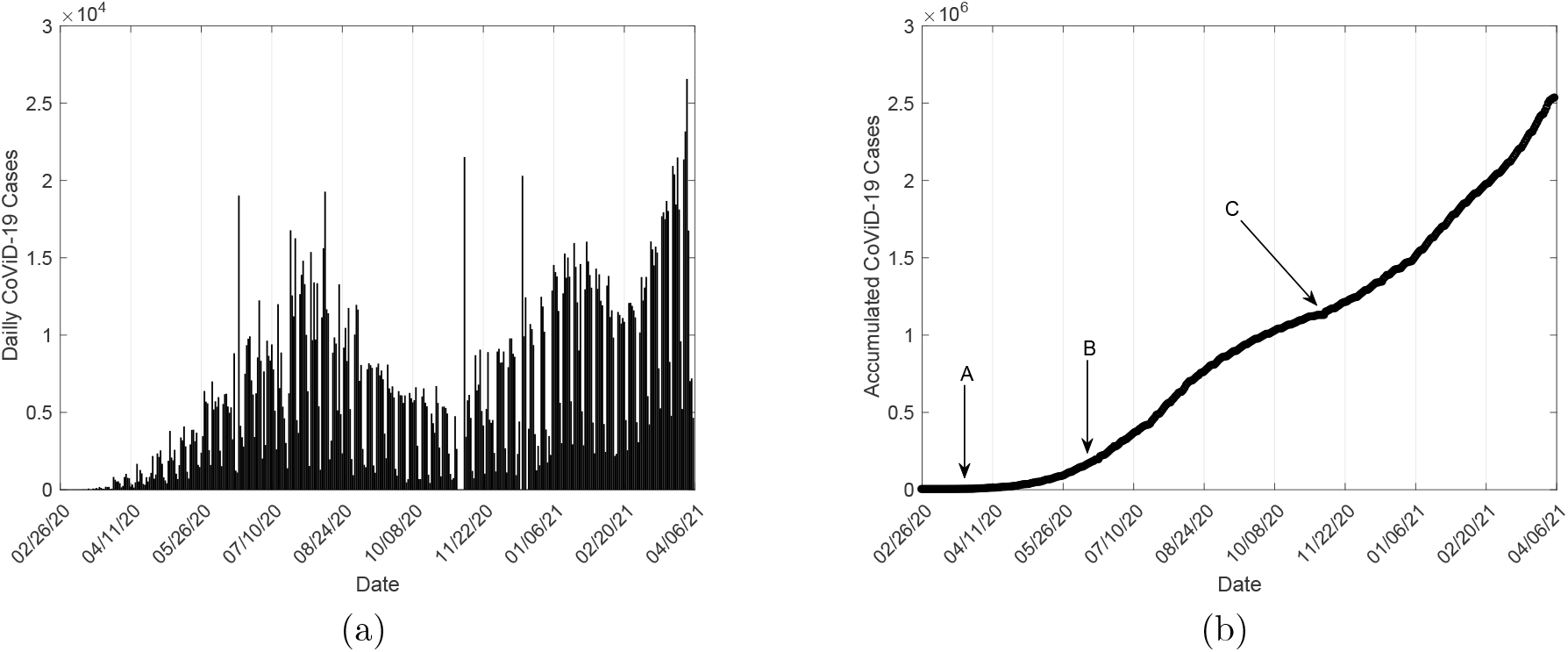
The daily (a) and accumulated (b) CoViD-19 cases collected from São Paulo State. In (b), *A* indicates the time at which isolation was introduced, *B* represents the time at which inflection point occurred, and *C* stands for the epidemic’s turn-over time.

In Figure B.1(b), *A* indicates the time at which isolation was introduced, and *B* represents the time at which the upward concavity changed to downward concavity in the first sigmoid (inflection point, which occurs when the daily cases reach the maximum value). In Saõ Paulo State, the isolation was introduced on March 24 (*A*), and the inflection point occurred after 80 *days* on June 12 (*B*). During this period almost all registered cases were severe CoviD-19. The downward concavity of the first sigmoid is broken on November 10 (*C*), and a second sigmoid initiates. Hence, *C* stands for the epidemic’s turn-over time, that is, the retaken of the epidemic.

Figure B.2 shows the daily (a) and accumulated (b) deaths due to CoViD-19.

**Figure B.2:**
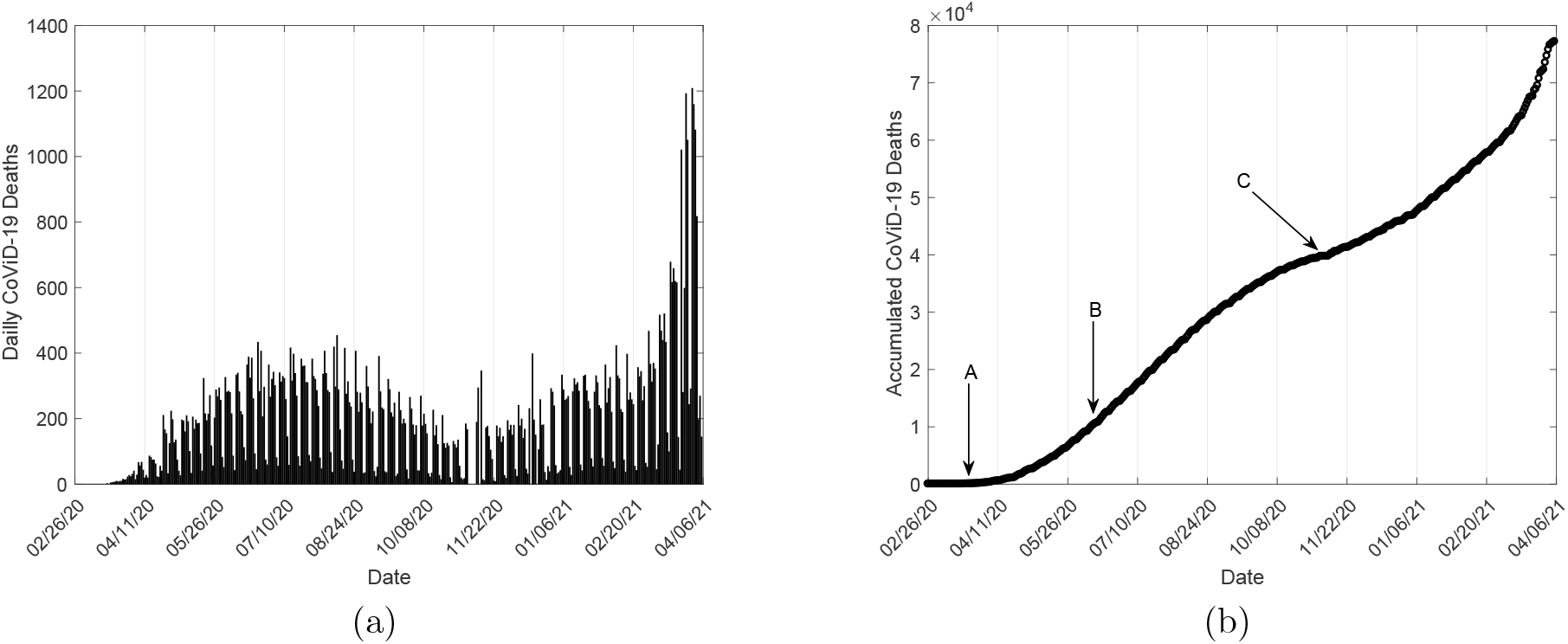
The daily (a) and accumulated (b) CoViD-19 deaths using data collected from São Paulo State.

The trend found in the CoViD-19 registered cases are observed in the accumulated deaths due to severe CoViD-19. Hence, we must retrieve the severe CoVid-19 from the observed deaths when quarantine ends and the relaxation begins.

Figure B.3 shows the population’s proportion in isolation [17]. Around the mean value *u* = 0.53 varies the daily isolation proportions during the quarantine.

**Figure B.3:**
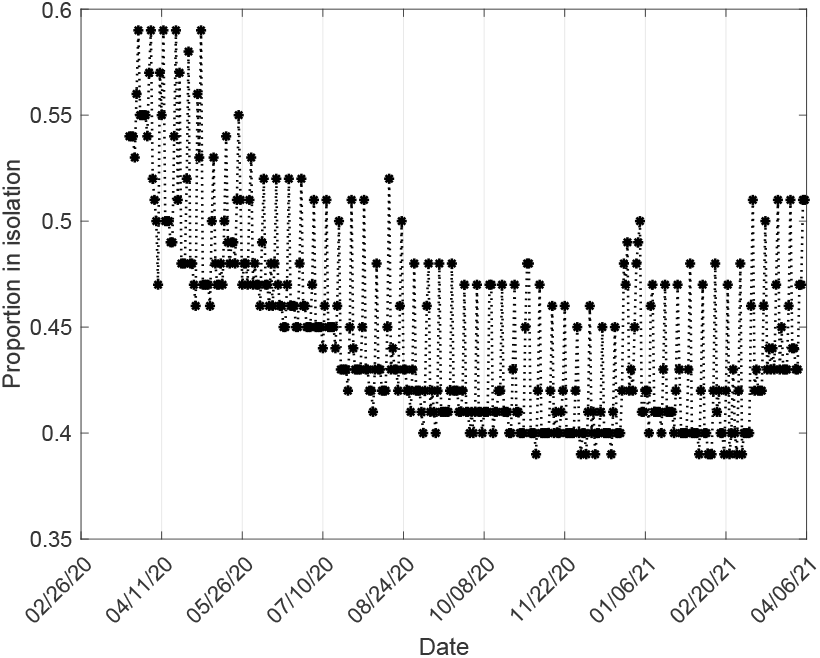
The proportion in isolation in São Paulo State.

The proportion in isolation decreases up to October, 2020. However, at the end of 2020 and beginning of 2021 we observe an increased isolation. This increase maybe due to the family gathering, which may increased the close contact among them.

## References

[1] Guirao A. The Covid-19 outbreak in Spain. A simple dynamics model, some lessons, and a theoretical framework for control response. Infectious Disease Modelling 2020; 5: 652–669. https://doi.org/10.1016/j.idm.2020.08.010.

[2] Battineni G, Chintalapudi N, Amenta F. SARS-CoV-2 epidemic calculation in Italy by SEIR compartmental models. Applied Computing and Informatics 2020; Emerald Publishing Limited: 11 pg. https://doi.org/10.1108/ACI-09-2020-0060.

[3] Yang HM, Lombardi Junior LP, Castro FFM, Yang AC. Mathematical model describing Covid-19 in São Paulo State, Brazil – Evaluating isolation as control mechanism and forecasting epidemiological scenarios of release. Epidemiology and Infection 2020 148: e155. doi: 10.1017/S0950268820001600.

[4] CDC, SARS-CoV-2 Variants. Updated Jan. 31, 2021. [accessed February 23, 2021]. Available from: https://www.cdc.gov/coronavirus/2019-ncov/cases-updates/variant-surveillance/variant-info.html.

[5] Yang HM, Lombardi Junior LP, Castro FFM, Campos AC. Mathematical modeling of the transmission of SARS-CoV-2 – Evaluating the impact of isolation in São Paulo State (Brazil) and lockdown in Spain associated with protective measures on the epidemic of covid-19. MedRxiv 2020. doi: https://doi.org/10.1101/2020.07.30.20165191.

[6] Yang HM, Lombardi Junior LP, Campos AC. Evaluating the trade-off between trans-missibility and virulence of SARS-CoV-2 by mathematical modeling. MedRxiv 2021. doi: https://doi.org/10.1101/2021.02.27.21252592.

[7] Sabino, EC, et al. Resurgence of COVID-19 in Manaus, Brazil, despite high seroprevalence. The Lancet 2021; 397 (10273): 452–455. doi: https://doi.org/10.1016/S0140-6736(21)00183-5.

[8] Korber B, et al. Tracking changes in SARS-CoV-2 spike: Evidence that D614G increases infectivity of the COVID-19 virus. Cell 2021; 182: 812-817. doi: https://doi.org/10.1016/j.cell.2020.06.043.

[9] SEADE, SP contra o novo coronavírus – Boletim completo. [accessed March 26, 2021]. Database [Internet]. Available from: https://www.seade.gov.br/coronavirus/?utm_source=portal&utm_medium=banner&utm_campaign=boletim-completo.

[10] Yang HM, Lombardi Junior LP, Campos AC. Are the SIR and SEIR models suitable to estimate the basic reproduction number for the CoViD-19 epidemic?. MedRxiv 2020. doi: https://doi.org/10.1101/2020.10.11.20210831.

[11] Informações COVID-19. [accessed March 26, 2021]. Database [Internet]. Available from: http://www.transparencia.am.gov.br/covid-19/monitoramento-covid-19/.

[12] King AA, Celle’s MD, Magpantay FMG, Rohani P. Avoidable errors in the modelling of outbreaks of emerging pathogens, with special reference to Ebola. Proc. R. Soc. B 2015; 282: 20150347. doi: http://dx.doi.org/10.1098/rspb.2015.0347.

[13] Arons MM, et al. Presymptomatic SARS-CoV-2 infections and transmission in a skilled nursing facility. The New Engl. Jour. Medicine 2020; April 24, 2020. doi: 10.1056/NEJ-Moa2008457.

[14] Anderson RM, May, RM. Infectious Diseases of Human. Dynamics and Control. Oxford, New York, Tokyo: Oxford University Press; 1991: 757 p.

[15] Yang HM. The basic reproduction number obtained from Jacobian and next generation matrices – A case study of dengue transmission modelling. BioSystems 2014; 126: 52–75.

[16] Yang HM, Greenhalgh D. Proof of conjecture in: The basic reproduction number obtained from Jacobian and next generation matrices – A case study of dengue transmission modelling. Appl. Math. Comput. 2015; 265: 103–107.

[17] Adesão ao isolamento social em SP; 2020 [accessed March 26, 2021]. Database [Internet]. Available from: https://www.saopaulo.sp.gov.br/coronavirus/isolamento.

